# Prevalence of Nonsuppressed Viral Load and Associated Factors Among Adults Receiving Antiretroviral Therapy in Eswatini, Lesotho, Malawi, Zambia, and Zimbabwe (2015-2017): Results from Population-Based Nationally-Representative Surveys

**DOI:** 10.1101/2020.07.13.20152553

**Authors:** Andreas D. Haas, Elizabeth Radin, Avi J. Hakim, Andreas Jahn, Neena Philip, Sasi Jonnalagadda, Suzue Saito, Andrea Low, Hetal Patel, Amee Schwitters, John Rogers, Koen Frederix, Evelyn Kim, George Bello, Daniel B Williams, Bharat Parekh, Karampreet Sachathep, Danielle T. Barradas, Thokozani Kalua, Sehin Birhanu, Godfrey Musuka, Owen Mugurungi, Beth A. Tippett Barr, Katrina Sleeman, Lloyd B Mulenga, Kyaw Thin, Trong Ao, Kristin Brown, Andrew C. Voetsch, Jessica J. Justman

**Affiliations:** ICAP-Columbia University, New York, NY, USA; Institute of Social and Preventive Medicine (ISPM), University of Bern, Bern, Switzerland; Division of Global HIV and TB, Center for Global Health, CDC, Atlanta, GA, USA; Ministry of Health Malawi, Lilongwe, Malawi; Division of Global HIV and TB, Center for Global Health, CDC Lesotho, Maseru, Lesotho; Division of Global HIV and TB, Center for Global Health, CDC Zimbabwe, Harare, Zimbabwe; Division of Global HIV and TB, Center for Global Health, CDC Malawi, Lilongwe, Malawi; Division of Global HIV and TB, Center for Global Health, CDC Zambia, Lusaka, Zambia; Ministry of Health & Child Welfare, Harare, Zimbabwe; Ministry of Health, Lusaka, Zambia; Research Coordination Unit, Ministry of Health, Maseru, Lesotho; Division of Global HIV and TB, Center for Global Health, CDC Eswatini, Mbabane, Eswatini

**Keywords:** ARV, viral load suppression, 90-90-90 targets, second-line ART, LMIC, sub-Saharan Africa

## Abstract

**Introduction:** The Joint United Nations Programme on HIV/AIDS (UNAIDS) has set a target of ≥90% of people living with HIV (PLHIV) receiving antiretroviral therapy (ART) to have viral load suppression (VLS). We examined factors associated with nonsuppressed viral Load (NVL).

**Methods:** We included PLHIV receiving ART aged 15–59 years from Eswatini, Lesotho, Malawi, Zambia, and Zimbabwe. Blood samples from PLHIV were analyzed for HIV RNA and recent exposure to antiretroviral drugs (ARVs). Outcomes were NVL (viral load ≥1000 copies/mL), virologic failure (VF; ARVs present and viral load ≥1000 copies/mL), interrupted ART (ARVs absent and viral load ≥1000 copies/mL), and receiving second-line ART. We calculated odds ratios and incidence rate ratios for factors associated with NVL, VF, interrupted ART, and switching to second-line ART.

**Results:** The prevalence of NVL was 11.2%: 8.2% experienced VF, and 3.0% interrupted ART. Younger age, male gender, less education, suboptimal adherence, receiving nevirapine, HIV non-disclosure, never having married, and residing in Zimbabwe, Lesotho, or Zambia were associated with higher odds of NVL. Among people with NVL, marriage, female gender, shorter ART duration, higher CD4 count, and alcohol use were associated with higher odds for interrupted ART and lower odds for VF. Many people with VF (44.8%) had CD4 counts <200 cells/µL, but few (0.31% per year) switched to second-line ART.

**Conclusions:** Countries are approaching UNAIDS VLS targets for adults. Treatment support for people initiating ART with asymptomatic HIV infection, scale-up of viral load monitoring, and optimized ART regimens may further reduce NVL prevalence.

## Introduction

Antiretroviral therapy (ART) interferes with viral replication and reduces the concentration of HIV RNA in the blood (viral load [VL]). VL suppression (VLS) is the most direct measure for the effectiveness of ART. Based on Joint United Nations Programme on HIV/AIDS (UNAIDS) modeled estimates, in 2017, 79% of PLHIV receiving ART in Eastern and Southern Africa had VLS [1]. UNAIDS has set a target for 90% of patients on ART to have VLS by 2020 [2]. Maintaining VLS keeps HIV-positive patients healthy and reduces the risk of HIV transmission [3–6]. Consistent adherence to an effective ART regimen is essential for maintaining VLS, but inadequate adherence and use of ineffective ART regimens are common, leading to reduced individual and public health benefits of ART [7,8].

The World Health Organization (WHO) recommends annual routine VL monitoring for patients receiving ART to detect adherence problems and treatment failure early, facilitating timely adherence support and ART regimen changes [9]. Recently, VL monitoring has been scaled up in resource-limited settings; however, in most sub-Saharan African countries, access remains limited. The proportion of patients who had received at least one VL test by mid-2016 was 19% in Malawi, 11% in Côte d’Ivoire, 49% in Kenya, 43% in Namibia, 9% in Tanzania, and 22% in Uganda [10]. For settings with limited VL testing capacity, WHO recommends targeted VL testing of patients with suspected treatment failure [9]. In settings without access to VL testing, health care workers rely on immunological or clinical criteria to detect ART failure [7]. The sensitivity and positive predictive value of immunological or clinical criteria for identifying patients with virologic failure (VF) are poor, and patients may not receive effective adherence support, may remain on ineffective ART regimens, or may switch to second-line ART unnecessarily [7]. Even where VL testing has been implemented, patients may not benefit from clinical management if results are not delivered in a timely manner or are not used for clinical decision making [7,11].

We analyzed nationally representative data from the population-based HIV impact assessment (PHIA) surveys to identify factors associated with nonsuppressed VL (NVL), VF, interrupted ART, and switching to second-line ART among adults in Lesotho, Malawi, Eswatini, Zambia, and Zimbabwe.

## Methods

### Survey design

The PHIA surveys are nationally representative cross-sectional household surveys conducted in 13 African countries and Haiti. PHIA surveys use a two-stage cluster-based sampling design. The surveys measure HIV incidence, HIV prevalence, and VLS prevalence among PLHIV and progress toward each of the UNAIDS 90-90-90 targets.

We used structured questionnaires to collect data on sociodemographic and behavioral characteristics. Survey participants were offered home-based HIV counseling and testing using each country’s national rapid testing algorithm with immediate return of results. HIV-positive participants who were not receiving ART were referred for treatment. HIV-positive blood specimens were tested for plasma VL. If an an insufficient volume of plasma was collected HIV VL testing was performed on dried blood spots (DBS). Positive specimens were also tested for CD4 cell counts, and for the presence of selected antiretroviral drugs (ARVs). In each country, samples were tested for the first-line ARVs efavirenz (EFV) and nevirapine (NVP) and the second-line ARV lopinavir (LPV). Malawi and Zambia also tested samples for the second-line ARV atazanavir (ATV). The survey was conducted between 2015 and 2017 in the five countries included in our analysis.

### Eligibility

We included HIV-positive adults aged 15–59 years who participated in the five PHIA surveys, who consented to biomarker testing, and who either reported current ART use or had detectable blood levels of selected ARVs. We excluded people who reported receiving ART for <4 months to account for people with high baseline VL who may not have had VLS at the time of data collection (<u>Figure 1</u>).

**Figure 1:**
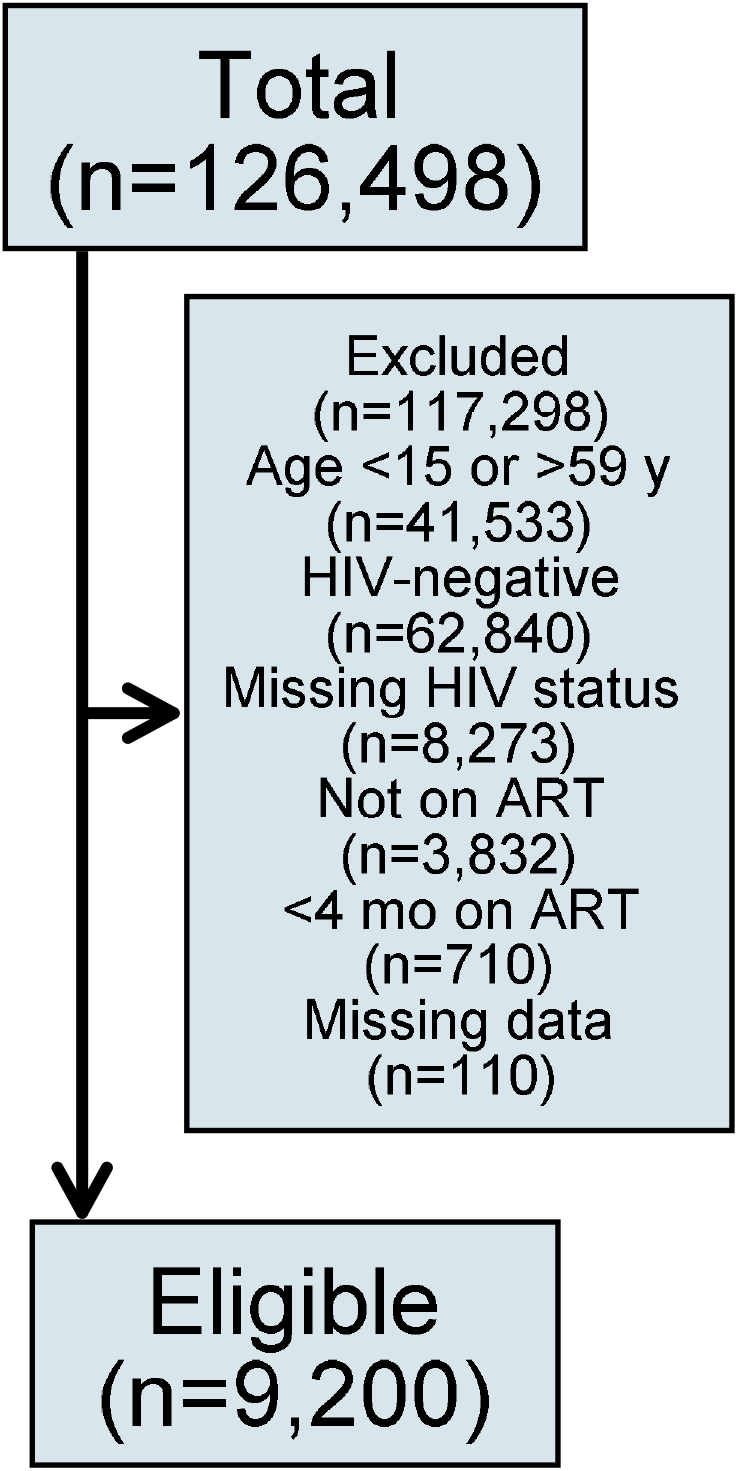
Flow diagram of HIV-positive adults aged 15–59 years on antiretroviral therapy (ART) who participated in the Population-based HIV Impact Assessment (PHIA) survey in Eswatini, Lesotho, Malawi, Zambia, and Zimbabwe (2015–2017) Data are unweighted numbers of participants. We excluded patients with missing self-reported data on current ART use or missing viral load or ARV detection testing results. Abbreviations: Y, years; mo, months.

### Definitions

We defined NVL as VL ≥1000 copies/mL because WHO uses this threshold to define treatment failure [9]. Patients with NVL were classified as either experiencing VF if any single ARV was detected in blood samples or as having interrupted ART if no ARVs were detected. We classified regimens as EFV-based if EFV was detected, as NVP-based if NVP was detected, and as protease inhibitor (PI)-based if either LPV or ATV was detected. We classified non-nucleoside reverse-transcriptase inhibitors (NNRTI)-based regimens as first-line ART and PI-based regimens as second-line ART [9]. CD4 cell count was categorized as <100 cells/µL, 100–199 cells/µL, 200–349 cells/µL, 350–499 cells/µL, and ≥500 cells/µL. VL was categorized as <40 copies/ml, 40-999 copies/ml and ≥1000 copies/ml. DBS VL samples could not be categorized and were excluded from this analysis.

We calculated ART duration as the date of the survey interview minus the self-reported ART start date. We categorized ART duration as <1, 1–2, 2–5, 6–10, and >10 years. We defined adherence to ART, based on self-reported data, as optimal if a participant reported missing in less than two doses in the last 30 days or suboptimal if the patient missed more than two doses in the last 30 days.

We used asset-based wealth index and categorized the wealth index scores into quintiles [12]. Households that received cash transfer, assistance for school fees, material support for education, or income generation support in cash or other forms in the last 12 months were considered to have received economic support. Work for which individuals received a salary, cash, or in-kind payment in the last 12 months was classified as paid work.

Alcohol use was assessed with the AUDIT-C three-item questionnaire. We used the cut-off AUDIT-C score of ≥4 in men and ≥3 in women to classify people screening positive for hazardous drinking [13,14].

### Statistical analysis

We used weighted summary statistics to describe characteristics of participants receiving ART and characteristics of participants with NVL, stratified by country. We used bivariate and multivariable logistic regression to calculate odds ratios (OR) to identify factors associated with NVL, interrupted ART, and VF. Logistic regression models accounted for stratification and clustering in the sample design. We estimated variance using the Taylor series linearization method. We used purposeful selection of covariates to build our final regression models [15].

We considered characteristics of ART and sociodemographic and behavioral characteristics in bivariate analysis. Variables associated with the outcome at a significance level of <0.20 in bivariate analysis [16,17] and variables that had been reported as important determinants of VLS or interruption of ART (i.e., wealth quintile, education, and ART duration) [18–24] were considered in multivariable analysis. We did not consider CD4 count and evidence of recent exposure to ARVs in multivariable analysis of factors associated with NVL because, like NVL, these are markers of treatment response or treatment adherence. Variables not significant at the level <0.05 in multivariable analysis were successively eliminated from multivariable models starting with the least significant variable. After eliminating a variable from the model, we compared the estimates of all coefficients that remained in the smaller model to their respective estimates from the larger model and added the eliminated variable back into the model if an estimate changed by more than 20%. We checked for interactions between independent variables that remained in the final model. We imputed missing data for all variables that were considered in multivariable analysis using multiple imputation with chained equation, ran multivariable analysis on 25 imputed datasets, and pooled results using Rubin’s rules [25–27]. Imputation models included all variables that were considered in multivariable analysis, NVL, VF, interruption of ART, CD4 cell count, region, and survey weights [27,28].

We calculated rates of switching to second-line ART as the number of patients who switched divided by the person years on first-line ART. We had no data on the timing of switching to second-line ART but assumed that patients switched after 1.85 years—the median time from initiation of first-line ART to switching to second-line ART in programs in resource-limited settings without access to routine VL monitoring [29]. We calculated incidence rate ratios (IRR) comparing switching rates by country, age, gender, CD4 count, and urban or rural residence using multivariable Poisson regression. In sensitivity analyses, we assumed that patients switched at the earliest and latest possible times (i.e., 6 months after initiating first-line ART and on the day of the survey). We excluded patients with unknown ART start date and unknown ART regimen from analysis of switching rates and factors associated with switching. To ensure national population-level representativeness of the survey, all analyses were weighted for selection probability, non-response, and non-coverage using survey weights. We report weighted percentages and unweighted numbers of participants in all analyses if not stated otherwise. Statistical analysis was done using Stata (Version 15, Stata Corporation, College Station, TX, USA).

### Ethical considerations

Local ethics committees and Institutional Review Boards approved the PHIA surveys. All participants provided written informed consent. Anonymized data were used for statistical analyses. More details on laboratory methods and research ethics are given in the appendix (<u>Text S1-S2</u>).

## Results

### Characteristics of participants

We included data from 9,200 people who tested HIV-positive in the survey: 2,177 from Lesotho, 1,406 from Malawi, 2,025 from Eswatini, 1,424 from Zambia, and 2,168 from Zimbabwe. Most were women (64.2%) and resided in rural areas (59.5%). Median age was 39 years (interquartile range [IQR], 32–46 years). Most (62.9%) were aged 25–44 years; about one-third (29.3%) were aged 45–59 years, 4.6% were aged 20–24 years, and 3.1% were aged 15–19 years. Most (61.5%) were married or cohabitating. Few (6.9%) reported being unaware of their HIV-positive status or being HIV-negative. Most (69.0%) had disclosed their HIV status to a family member (<u>Table 1</u>). The prevalence of people who disclosed their HIV status is shown in <u>Table S1</u>. The prevalence of people who screened positive for problematic or hazardous drinking or active alcohol use disorder was 8.3% (<u>Table S2</u>).

**Table 1:** Sociodemographic characteristics of HIV-positive adults receiving antiretroviral therapy in Lesotho, Malawi, Eswatini, Zambia and Zimbabwe (2015–2017)

| Characteristic | Lesotho<br>N=2,177 | Malawi<br>N=1,406 | Eswatini<br>N=2,025 | Zambia<br>N=1,424 | Zimbabwe<br>N=2,168 | Total<br>N=9,200 |
| --- | --- | --- | --- | --- | --- | --- |
| Residence, n (%) | 2,177 (100.0) | 1,406 (100.0) | 2,025 (100.0) | 1,424 (100.0) | 2,168 (100.0) | 9,200 (100.0) |
| Urban | 991 (49.8) | 644 (24.9) | 487 (29.1) | 868 (62.3) | 656 (35.8) | 3,646 (40.5) |
| Rural | 1,186 (50.2) | 762 (75.1) | 1,538 (70.9) | 556 (37.7) | 1,512 (64.2) | 5,554 (59.5) |
| Gender, n (%) | 2,177 (100.0) | 1,406 (100.0) | 2,025 (100.0) | 1,424 (100.0) | 2,168 (100.0) | 9,200 (100.0) |
| Male | 646 (38.0) | 401 (35.6) | 587 (30.3) | 414 (35.8) | 660 (36.3) | 2,708 (35.8) |
| Female | 1,531 (62.0) | 1,005 (64.4) | 1,438 (69.7) | 1,010 (64.2) | 1,508 (63.7) | 6,492 (64.2) |
| Age, years n (%) | 2,177 (100.0) | 1,406 (100.0) | 2,025 (100.0) | 1,424 (100.0) | 2,168 (100.0) | 9,200 (100.0) |
| 15-19 | 56 (2.7) | 22 (2.0) | 70 (3.3) | 29 (2.6) | 83 (4.6) | 260 (3.1) |
| 20-24 | 116 (4.7) | 76 (4.3) | 124 (6.1) | 65 (4.9) | 85 (4.4) | 466 (4.6) |
| 25-44 | 1,293 (61.6) | 923 (65.0) | 1,294 (67.0) | 895 (62.2) | 1,287 (61.4) | 5,692 (62.9) |
| 45-59 | 712 (30.9) | 385 (28.7) | 537 (23.6) | 435 (30.3) | 713 (29.7) | 2,782 (29.3) |
| Median (IQR) | 39 (32-47) | 39 (32-45) | 37 (30-44) | 40 (33-46) | 39 (32-46) | 39 (32-46) |
| Wealth quintile, n (%) | 2,174 (100.0) | 1,406 (100.0) | 2,024 (100.0) | 1,413 (100.0) | 2,168 (100.0) | 9,185 (100.0) |
| Lowest | 440 (17.4) | 147 (14.1) | 512 (23.1) | 119 (8.2) | 554 (21.1) | 1,772 (15.8) |
| Second | 493 (21.3) | 172 (17.0) | 457 (21.7) | 157 (10.8) | 436 (18.6) | 1,715 (16.7) |
| Middle | 480 (22.5) | 193 (18.3) | 429 (20.7) | 295 (20.2) | 399 (19.2) | 1,796 (19.6) |
| Fourth | 400 (20.0) | 287 (23.4) | 349 (19.2) | 371 (26.0) | 393 (21.7) | 1,800 (22.9) |
| Highest | 361 (18.8) | 607 (27.1) | 277 (15.3) | 471 (34.8) | 386 (19.3) | 2,102 (24.9) |
| Education, n (%) | 2,176 (100.0) | 1,405 (100.0) | 2,022 (100.0) | 1,424 (100.0) | 2,168 (100.0) | 9,195 (100.0) |
| No or primary | 1,309 (59.1) | 999 (77.5) | 890 (41.3) | 649 (44.0) | 891 (36.9) | 4,738 (51.7) |
| Secondary or higher | 867 (40.9) | 406 (22.5) | 1,132 (58.7) | 775 (56.0) | 1,277 (63.1) | 4,457 (48.3) |
| Marital status, n (%) | 2,171 (100.0) | 1,404 (100.0) | 2,012 (100.0) | 1,420 (100.0) | 2,163 (100.0) | 9,170 (100.0) |
| Never married | 330 (15.5) | 76 (4.6) | 654 (33.0) | 133 (10.2) | 208 (10.0) | 1,401 (10.6) |
| Married/living together | 1,130 (54.4) | 909 (65.9) | 1,019 (50.5) | 873 (63.1) | 1,287 (61.0) | 5,218 (61.5) |
| Divorced, separated or widowed | 711 (30.1) | 419 (29.5) | 339 (16.5) | 414 (26.7) | 668 (29.0) | 2,551 (27.9) |
| Economic support, n (%) | 2,177 (100.0) | 1,406 (100.0) | 2,025 (100.0) | 1,424 (100.0) | 2,168 (100.0) | 9,200 (100.0) |
| No | 1,720 (80.4) | 1,302 (91.1) | 1,204 (60.7) | 1,364 (95.8) | 1,871 (87.1) | 7,461 (88.1) |
| Yes | 457 (19.6) | 104 (8.9) | 821 (39.3) | 60 (4.2) | 297 (12.9) | 1,739 (11.9) |
| Paid work, n (%) | 2,177 (100.0) | 1,406 (100.0) | 2,025 (100.0) | 1,421 (100.0) | 2,167 (100.0) | 9,196 (100.0) |
| Yes | 871 (43.9) | 480 (32.8) | 936 (48.5) | 537 (40.2) | 763 (37.4) | 3,587 (38.2) |
| No | 1,306 (56.1) | 926 (67.2) | 1,089 (51.5) | 884 (59.8) | 1,404 (62.6) | 5,609 (61.8) |
| Disclosure to a family member, n (%) | 2,177 (100.0) | 1,406 (100.0) | 2,025 (100.0) | 1,424 (100.0) | 2,168 (100.0) | 9,200 (100.0) |
| No | 540 (25.9) | 344 (24.2) | 660 (32.6) | 291 (20.5) | 507 (24.9) | 2,342 (24.2) |
| Yes | 1,525 (68.1) | 977 (69.4) | 1,307 (64.1) | 1,005 (70.1) | 1,544 (68.8) | 6,358 (69.0) |
| Unknown (unaware of HIV status) | 112 (5.9) | 85 (6.4) | 58 (3.3) | 128 (9.3) | 117 (6.3) | 500 (6.9) |

Abbreviations: N, Number of eligible participants; n, number of participants providing a valid response (weighted %). IQR, interquartile range.

### Characteristics of and response to ART

Median duration on ART was reported as 4 years (IQR, 2–7 years). On the basis of ARV testing, most patients (91.3%) received an EFV-based first-line regimen. The proportion of patients who received second-line regimens containing PIs ranged from 0.3% in Zimbabwe to 3.4% in Eswatini. Of the patients, 90.6% reported optimal adherence. The median CD4 count was 467 cells/µL (IQR, 315–641 cells/µL). Overall, 20.5% (1,837/8,972) of people who received a plasma viral load test had a detectable VL of ≥40 copies/mL. The prevalence NVL was 11.2% (n=964): 8.2% (n=686) had evidence of VF and 3.0% (n=278) of interrupted ART (<u>Table 2</u>).

**Table 2:**
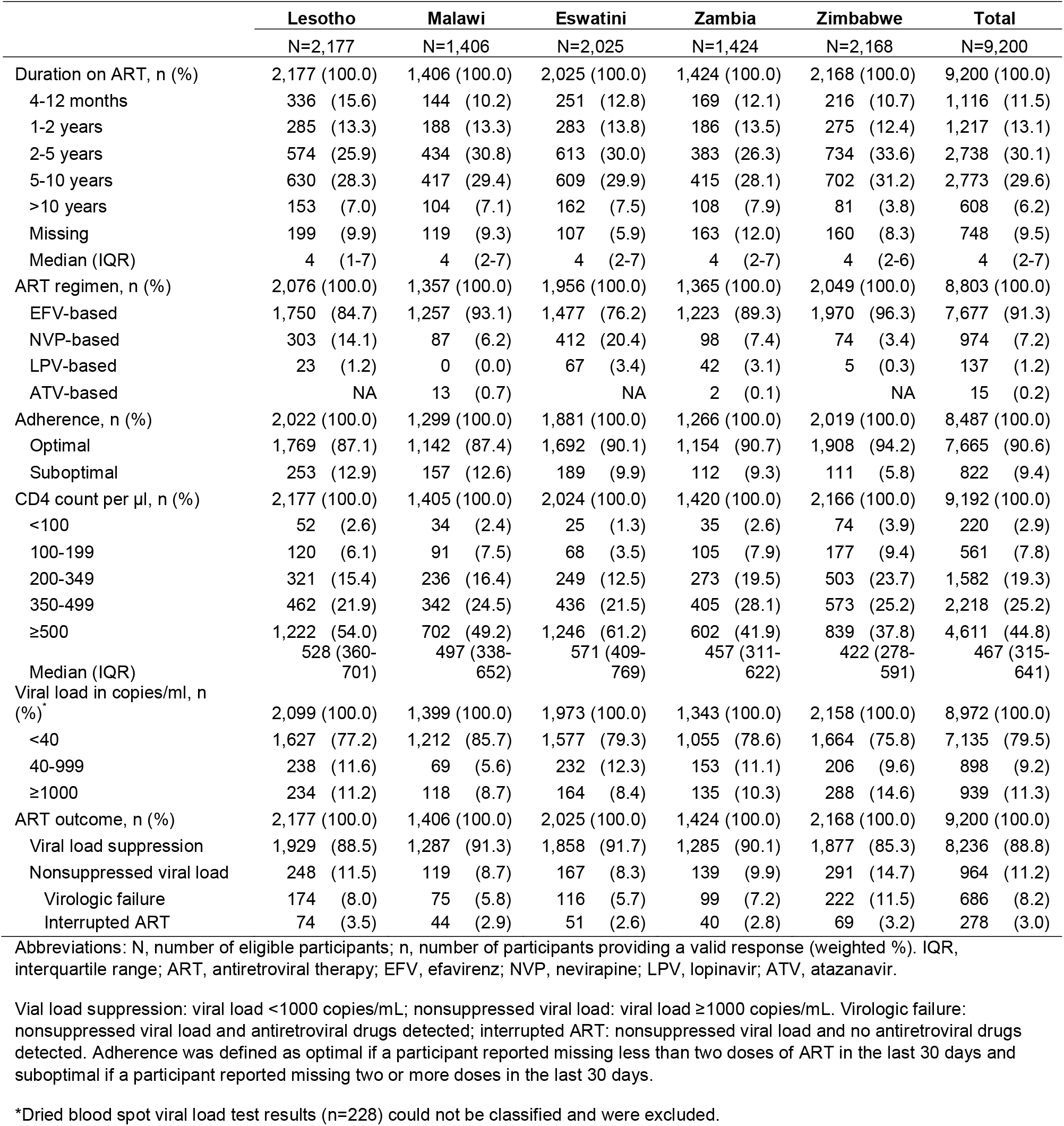
Characteristics of and response to antiretroviral therapy (ART) among HIV-positive adults in Lesotho, Malawi, Eswatini, Zambia and Zimbabwe (2015-2017)

### Characteristics of participants with NVL

<u>Table 3</u> shows the characteristics of participants with NVL. One-quarter of patients with NVL (26.7%) had evidence of interrupted ART, and three-quarters (73.3%) were experiencing VF. Most patients with NVL (59.8%) were women. Median age of patients with NVL was 36 years (IQR, 29–43 years). Median CD4 count among patients with VF was 222 cells/µL (IQR, 116–381 cells/µL), and median duration on ART was 4.5 years (IQR, 2.6–6.9). Only 1.8% of patients with VF received second-line ART.

**Table 3:**
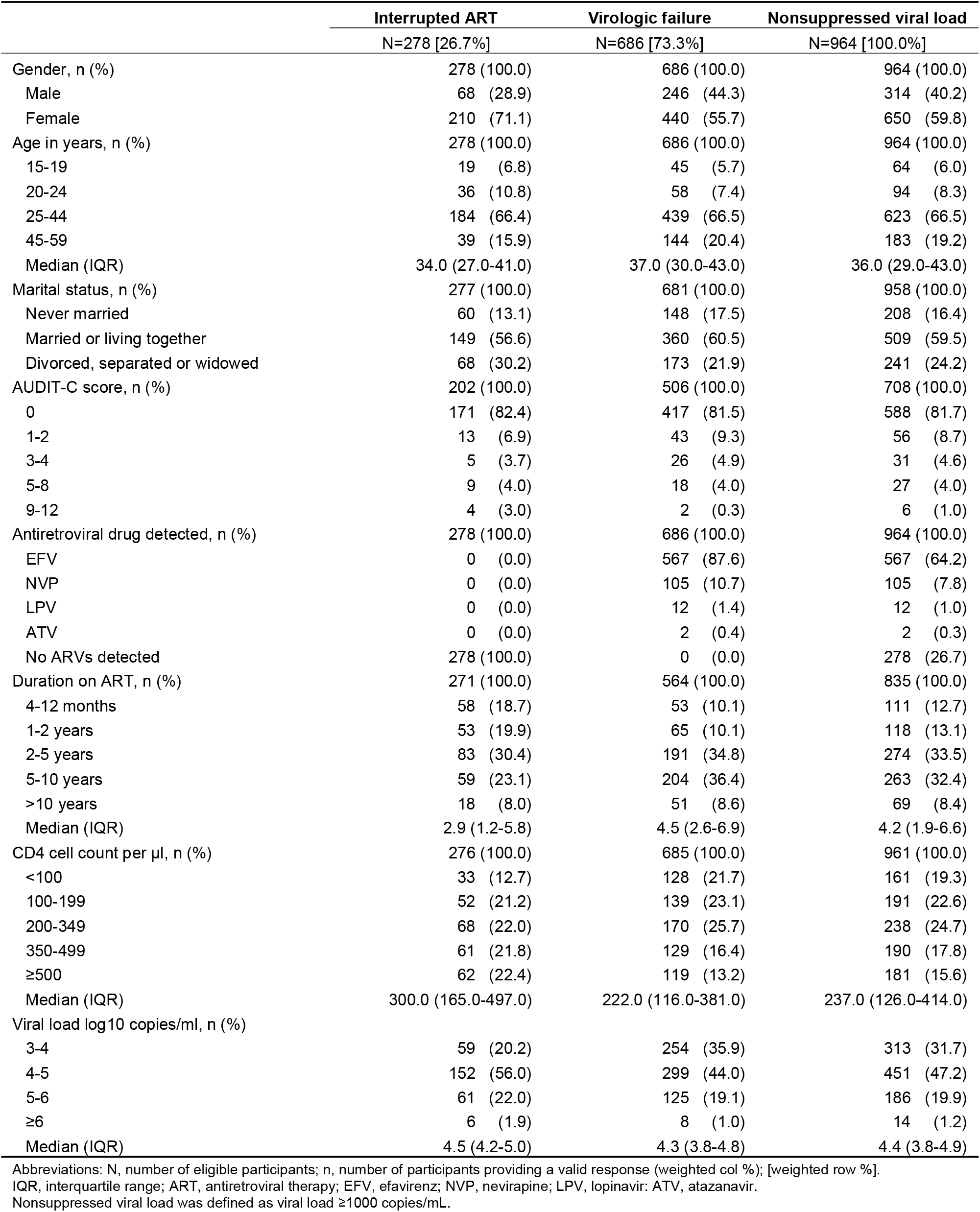
**Characteristics of adults on antiretroviral therapy with nonsuppressed viral load: Results from the Lesotho, Malawi, Eswatini, Zambia, and Zimbabwe Population-based HIV Impact Assessment survey (2015–2017)**

|  | Interrupted ART | Virologic failure | Nonsuppressed viral load |
| --- | --- | --- | --- |
|  | N=278 [26.7%] | N=686 [73.3%] | N=964 [100.0%] |
| Gender, n (%) | 278 (100.0) | 686 (100.0) | 964 (100.0) |
| Male | 68 (28.9) | 246 (44.3) | 314 (40.2) |
| Female | 210 (71.1) | 440 (55.7) | 650 (59.8) |
| Age in years, n (%) | 278 (100.0) | 686 (100.0) | 964 (100.0) |
| 15-19 | 19 (6.8) | 45 (5.7) | 64 (6.0) |
| 20-24 | 36 (10.8) | 58 (7.4) | 94 (8.3) |
| 25-44 | 184 (66.4) | 439 (66.5) | 623 (66.5) |
| 45-59 | 39 (15.9) | 144 (20.4) | 183 (19.2) |
| Median (IQR) | 34.0 (27.0-41.0) | 37.0 (30.0-43.0) | 36.0 (29.0-43.0) |
| Marital status, n (%) | 277 (100.0) | 681 (100.0) | 958 (100.0) |
| Never married | 60 (13.1) | 148 (17.5) | 208 (16.4) |
| Married or living together | 149 (56.6) | 360 (60.5) | 509 (59.5) |
| Divorced, separated or widowed | 68 (30.2) | 173 (21.9) | 241 (24.2) |
| AUDIT-C score, n (%) | 202 (100.0) | 506 (100.0) | 708 (100.0) |
| 0 | 171 (82.4) | 417 (81.5) | 588 (81.7) |
| 1-2 | 13 (6.9) | 43 (9.3) | 56 (8.7) |
| 3-4 | 5 (3.7) | 26 (4.9) | 31 (4.6) |
| 5-8 | 9 (4.0) | 18 (4.0) | 27 (4.0) |
| 9-12 | 4 (3.0) | 2 (0.3) | 6 (1.0) |
| Antiretroviral drug detected, n (%) | 278 (100.0) | 686 (100.0) | 964 (100.0) |
| EFV | 0 (0.0) | 567 (87.6) | 567 (64.2) |
| NVP | 0 (0.0) | 105 (10.7) | 105 (7.8) |
| LPV | 0 (0.0) | 12 (1.4) | 12 (1.0) |
| ATV | 0 (0.0) | 2 (0.4) | 2 (0.3) |
| No ARVs detected | 278 (100.0) | 0 (0.0) | 278 (26.7) |
| Duration on ART, n (%) | 271 (100.0) | 564 (100.0) | 835 (100.0) |
| 4-12 months | 58 (18.7) | 53 (10.1) | 111 (12.7) |
| 1-2 years | 53 (19.9) | 65 (10.1) | 118 (13.1) |
| 2-5 years | 83 (30.4) | 191 (34.8) | 274 (33.5) |
| 5-10 years | 59 (23.1) | 204 (36.4) | 263 (32.4) |
| >10 years | 18 (8.0) | 51 (8.6) | 69 (8.4) |
| Median (IQR) | 2.9 (1.2-5.8) | 4.5 (2.6-6.9) | 4.2 (1.9-6.6) |
| CD4 cell count per µl, n (%) | 276 (100.0) | 685 (100.0) | 961 (100.0) |
| <100 | 33 (12.7) | 128 (21.7) | 161 (19.3) |
| 100-199 | 52 (21.2) | 139 (23.1) | 191 (22.6) |
| 200-349 | 68 (22.0) | 170 (25.7) | 238 (24.7) |
| 350-499 | 61 (21.8) | 129 (16.4) | 190 (17.8) |
| ≥500 | 62 (22.4) | 119 (13.2) | 181 (15.6) |
| Median (IQR) | 300.0 (165.0-497.0) | 222.0 (116.0-381.0) | 237.0 (126.0-414.0) |
| Viral load log<sub>10</sub> copies/ml, n (%) |  |  |  |
| 3-4 | 59 (20.2) | 254 (35.9) | 313 (31.7) |
| 4-5 | 152 (56.0) | 299 (44.0) | 451 (47.2) |
| 5-6 | 61 (22.0) | 125 (19.1) | 186 (19.9) |
| ≥6 | 6 (1.9) | 8 (1.0) | 14 (1.2) |
| Median (IQR) | 4.5 (4.2-5.0) | 4.3 (3.8-4.8) | 4.4 (3.8-4.9) |

Abbreviations: N, number of eligible participants; n, number of participants providing a valid response (weighted col %); [weighted row %]. IQR, interquartile range; ART, antiretroviral therapy; EFV, efavirenz; NVP, nevirapine; LPV, lopinavir; ATV, atazanavir. Nonsuppressed viral load was defined as viral load ≥1000 copies/mL.

### Factors associated with NVL

The adjusted odds ratios (aOR) for NVL for HIV-positive patients in Zimbabwe (aOR, 2.61; 95% confidence interval [CI]: 2.03–3.35), Lesotho (aOR, 1.61; 95% CI: 1.26–2.05), and Zambia (aOR, 1.52; 95% CI: 1.14–2.02) were higher than in Eswatini. The odds for NVL in HIV-positive patients aged 15–24 years were 2–3 times higher than in those aged 45–59 years. The odds of NVL were slightly increased in men (aOR, 1.28; 95% CI: 1.05–1.55), in people with low educational levels (aOR, 1.27; 95% CI: 1.06–1.52), in those who had never married (aOR, 1.42; 95% CI: 1.02–1.98), and in participants who had not disclosed their HIV status to a family member (aOR, 1.32; 95% CI: 1.06–1.66). People receiving NVP-based ART had higher odds for NVL (aOR, 1.84; 95% CI: 1.34–2.51) than those receiving EFV-based regimens. Odds for NVL were also higher in patients who reported sub-optimal adherence (aOR, 1.81; 95% CI: 1.37–2.39; <u>Table 4</u>).

**Table 4:** Factors associated with nonsuppressed viral load among HIV-positive participants in the Population-based HIV Impact Assessment survey in Eswatini, Lesotho, Malawi, Zambia, and Zimbabwe (2015–2017)

|  | Bivariate analysis |  | Multivariable analysis |  |
| --- | --- | --- | --- | --- |
|  | OR (95% CI) | P-value | aOR (95% CI) | P-value |
| **Characteristics** |  |  |  |  |
| Country residence |  | <0.001 |  | <0.001 |
| Lesotho | 1.43 (1.14-1.79) |  | 1.61 (1.26-2.05) |  |
| Malawi | 1.05 (0.78-1.44) |  | 1.25 (0.90-1.74) |  |
| Eswatini | 1.00 |  | 1.00 |  |
| Zambia | 1.22 (0.94-1.57) |  | 1.52 (1.14-2.02) |  |
| Zimbabwe | 1.90 (1.53-2.35) |  | 2.61 (2.03-3.35) |  |
| Age, years |  | <0.001 |  | <0.001 |
| 15-19 | 3.47 (2.23-5.41) |  | 2.15 (1.23-3.78) |  |
| 20-24 | 3.17 (2.17-4.62) |  | 3.13 (2.08-4.71) |  |
| 25-44 | 1.70 (1.34-2.14) |  | 1.81 (1.42-2.32) |  |
| 45-59 | 1.00 |  | 1.00 |  |
| Gender |  | 0.022 |  | 0.013 |
| Male | 1.24 (1.03-1.48) |  | 1.28 (1.05-1.55) |  |
| Female | 1.00 |  | 1.00 |  |
| Marital status |  | <0.001 |  | 0.110 |
| Never married | 1.72 (1.33-2.23) |  | 1.42 (1.02-1.98) |  |
| Married or living together | 1.00 |  | 1.00 |  |
| Divorced, separated or widowed | 0.89 (0.72-1.09) |  | 1.02 (0.82-1.27) |  |
| Education |  | 0.977 |  | 0.010 |
| No or primary | 1.00 (0.83-1.20) |  | 1.27 (1.06-1.52) |  |
| Secondary or higher | 1.00 |  | 1.00 |  |
| Disclosure to a family member |  | 0.003 |  | 0.014 |
| No | 1.29 (1.04-1.61) |  | 1.32 (1.06-1.66) |  |
| Yes | 1.00 |  | 1.00 |  |
| Not self-reported positive | 1.64 (1.16-2.32) |  | 1.39 (0.96-2.02) |  |
| **Characteristics of antiretroviral therapy** |  |  |  |  |
| Antiretroviral regimen |  | 0.003 |  | 0.001 |
| EFV-based | 1.00 |  | 1.00 |  |
| NVP-based | 1.66 (1.23-2.24) |  | 1.84 (1.34-2.51) |  |
| PI-based | 1.65 (0.80-3.39) |  | 1.86 (0.88-3.89) |  |
| Adherence |  | <0.001 |  | <0.001 |
| Optimal | 1.00 |  | 1.00 |  |
| Suboptimal | 1.91 (1.46-2.50) |  | 1.81 (1.37-2.39) |  |

Abbreviations: ART, antiretroviral therapy; EFV, efavirenz; NVP, nevirapine; PI, protease inhibitor; OR, odds ratio; aOR, adjusted odds ratios. Multivariable analysis adjusted for country, age, gender, marital status, education, disclosure of HIV status to a family member, antiretroviral regimen, and adherence. Nonsuppressed viral load was defined as viral load $\geq 1000$ copies/mL. P-values of categorical variables are for joint test for significance.

### Factors associated with interrupted ART and VF among people with NVL

<u>Table 5</u> shows aORs comparing the odds for interrupted ART and VF among patients with NVL. The adjusted odds for interrupted ART among HIV-positive patients aged 15–19 years were 3 times (aOR, 3.04; 95% CI: 1.30–7.11) higher than in those aged 44–59 years. Women had almost twice the odds (aOR, 1.85; 95% CI: 1.01–3.38) of having interrupted ART than men. Married (aOR, 1.85; 95% CI: 1.07– 3.21) and divorced patients (aOR, 2.64; 95% CI: 1.40–4.89) had higher odds for interrupted ART than patients who never married. The odds for interrupted ART were about 3 times higher in patients receiving ART for <2 years than in people receiving ART for 5–10 years. Patients with a CD4 count of 350–500 cells/µL (aOR, 2.11; 95% CI: 1.19–3.74) and those with a CD4 count ≥500 cells/µL (aOR, 2.59; 95% CI: 1.26–5.30) had higher odds of interrupted ART than people with a CD4 cell count <100 cells/µL. Factors associated with interrupted ART were inversely associated with VF because people with NVL either had interrupted ART or VF (Table 5).

**Table 5:** Factors associated with interrupted ART and virologic failure among participants with nonsuppressed viral load in Eswatini, Lesotho, Malawi, Zambia, and Zimbabwe (2015–2017)

|  | Interrupted ART |  |  |  | Virologic failure |  |  |  |
| --- | --- | --- | --- | --- | --- | --- | --- | --- |
|  | Bivariate analysis |  | Multivariable analysis |  | Bivariate analysis |  | Multivariable analysis |  |
|  | OR (95% CI) | P-value | aOR (95% CI) | P-value | OR (95% CI) | P-value | aOR (95% CI) | P-value |
| **Sociodemographic characteristics** |  |  |  |  |  |  |  |  |
| Age in years |  | 0.250 |  | 0.056 |  | 0.250 |  | 0.056 |
| 15-19 | 1.52 (0.68-3.38) |  | 3.04 (1.30-7.11) |  | 0.66 (0.30-1.47) |  | 0.32 (0.14-0.76) |  |
| 20-24 | 1.88 (0.99-3.58) |  | 1.89 (0.88-4.07) |  | 0.53 (0.28-1.01) |  | 0.54 (0.24-1.18) |  |
| 25-44 | 1.28 (0.81-2.02) |  | 1.12 (0.71-1.78) |  | 0.78 (0.49-1.23) |  | 0.89 (0.56-1.42) |  |
| 45-59 | 1.00 |  | 1.00 |  | 1.00 |  | 1.00 |  |
| Gender |  | 0.002 |  | 0.047 |  | 0.002 |  | 0.047 |
| Male | 1.00 |  | 1.00 |  | 1.00 |  | 1.00 |  |
| Female | 1.96 (1.29-2.97) |  | 1.85 (1.01-3.38) |  | 0.51 (0.34-0.78) |  | 0.54 (0.29-0.99) |  |
| Marital status |  | 0.043 |  | 0.012 |  | 0.043 |  | 0.012 |
| Never married | 1.00 |  | 1.00 |  | 1.00 |  | 1.00 |  |
| Married or living together | 1.25 (0.83-1.89) |  | 1.85 (1.07-3.21) |  | 0.80 (0.53-1.20) |  | 0.53 (0.31-0.93) |  |
| Divorced, separated or widowed | 1.84 (1.14-2.98) |  | 2.64 (1.40-4.98) |  | 0.54 (0.34-0.88) |  | 0.37 (0.19-0.71) |  |
| **Antiretroviral therapy characteristics** |  |  |  |  |  |  |  |  |
| Duration on ART in years |  | 0.001 |  | <0.001 |  | 0.001 |  | <0.001 |
| <1 | 2.77 (1.47-5.24) |  | 3.13 (1.60-6.12) |  | 0.39 (0.20-0.76) |  | 0.34 (0.17-0.71) |  |
| 1-2 | 2.77 (1.61-4.78) |  | 3.24 (1.79-5.86) |  | 0.37 (0.21-0.65) |  | 0.32 (0.17-0.59) |  |
| 2-5 | 1.34 (0.85-2.11) |  | 1.31 (0.79-2.16) |  | 0.74 (0.47-1.17) |  | 0.75 (0.45-1.24) |  |
| 5-10 | 1.00 |  | 1.00 |  | 1.00 |  | 1.00 |  |
| >10 | 1.54 (0.76-3.12) |  | 1.73 (0.86-3.52) |  | 0.69 (0.33-1.40) |  | 0.62 (0.30-1.26) |  |
| CD4 cell count per µl |  | 0.004 |  | 0.048 |  | 0.004 |  | 0.048 |
| <100 | 1.00 |  | 1.00 |  | 1.00 |  | 1.00 |  |
| 100-199 | 1.58 (0.97-2.56) |  | 1.75 (1.09-2.82) |  | 0.65 (0.40-1.05) |  | 0.59 (0.37-0.95) |  |
| 200-349 | 1.47 (0.81-2.66) |  | 1.51 (0.84-2.73) |  | 0.69 (0.38-1.25) |  | 0.67 (0.37-1.21) |  |
| 350-499 | 2.27 (1.34-3.87) |  | 2.11 (1.19-3.74) |  | 0.44 (0.26-0.76) |  | 0.48 (0.27-0.85) |  |
| ≥500 | 2.90 (1.61-5.24) |  | 2.59 (1.26-5.30) |  | 0.35 (0.19-0.63) |  | 0.40 (0.19-0.81) |  |
| **Behavioral characteristics** |  |  |  |  |  |  |  |  |
| AUDIT-C score |  | <0.001 |  | 0.001 |  | <0.001 |  | 0.001 |
| 0 | 1.00 |  | 1.00 |  | 1.00 |  | 1.00 |  |
| 1-2 | 0.73 (0.36-1.51) |  | 0.97 (0.38-2.49) |  | 1.33 (0.67-2.63) |  | 1.01 (0.41-2.47) |  |
| 3-4 | 0.75 (0.47-1.21) |  | 1.00 (0.49-2.05) |  | 1.26 (0.76-2.11) |  | 0.93 (0.46-1.87) |  |
| 5-8 | 0.99 (0.54-1.79) |  | 1.84 (0.80-4.24) |  | 1.00 (0.55-1.81) |  | 0.55 (0.26-1.20) |  |
| 9-12 | 10.51 (3.53-31.30) |  | 12.64 (3.71-43.06) |  | 0.10 (0.04-0.28) |  | 0.09 (0.03-0.28) |  |

Abbreviations: ART, antiretroviral therapy; OR, odds ratio; aOR, adjusted odds ratios.
Nonsuppressed viral load was defined as viral load ≥1000 copies/mL. Patients with nonsuppressed viral load without detectable concentrations of antiretroviral medication in their blood were considered to have interrupted ART. Multivariable analysis adjusted for all variables shown in the table. P-values of categorical variables are for joint test for significance.

### Rates of and factors associated with switching to second-line ART

Overall, 131 (1.4%) of 8,066 participants with known date of ART initiation and known regimen switched to second-line ART, for an annual switching rate of 0.31% (95% CI: 0.25–0.40). Annual switching rates were 0.22% (95% CI: 0.14–0.36) in Lesotho, 0.15% (95% CI: 0.08–0.35) in Malawi, 0.72% (95% CI: 0.54– 0.96) in Eswatini, 0.73% (95% CI: 0.52–1.04) in Zambia, and 0.06% (95% CI: 0.02–0.33) in Zimbabwe. Compared to Eswatini, adjusted switching rates were more than 10 times lower in Zimbabwe (IRR, 0.07; 95% CI: 0.02–0.24), 5 times lower in Malawi (IRR, 0.19; 95% CI: 0.09–0.44), more than 3 times lower in Lesotho (IRR, 0.28; 95% CI: 0.15–0.52), and similar in Zambia (IRR, 0.86; 95% CI: 0.57–1.32).

People with CD4 counts <100 cells/µL were more than 3 times (IRR, 3.52; 95% CI: 1.05–11.76) and those with CD4 counts of 100–199 cells/µL were more than 2 times (IRR, 2.29; 95% CI: 1.05–5.01) as likely to switch to second-line ART than those with CD4 count of ≥500 cells/µL. Age, gender, and residence (rural or urban) were not associated with switching rates.

Analysis of switching rates was not sensitive to assumptions on timing of switching. The overall annual switching rate remained 0.31% whether we assumed that patients switched 6 months after initiation of first-line ART or on the day of the survey (Table S3).

## Discussion

These population-based nationally representative surveys showed that these five southern African countries are approaching UNAIDS VLS targets despite limited access to VL monitoring. We found that 11.2% of adults who reported current ART use or had detectable blood levels of ARVs had a VL of ≥1000 copies/mL. Sociodemographic characteristics, current ART regimen, adherence, and HIV status disclosure were associated with NVL. One-quarter of patients with NVL had no detectable blood concentrations of ARVs and were classified as having interrupted ART, while three-quarters of patients with NVL had evidence of recent ARV exposure and were classified as experiencing VF. Marriage, female gender, shorter ART duration, higher CD4 count, and alcohol use were associated with higher odds for interrupted ART and lower odds for VF. Almost half of the people with VF were severely immunosuppressed although they received ART for several years. Switching rates varied considerably between countries, but even in countries with the highest switching rates, very few patients had switched to second-line therapy.

Low switching rates, as documented in this study, indicate substantial gaps in monitoring of ART and/or clinical management of patients with treatment failure. HIV cohort studies from low-income and middle-income countries showed that each year about 3% of adults have a confirmed virological treatment failure of first-line ART and require switching to second-line ART [7,30]. We found nationally representative switching rates far below the annual failure rate of 3%: annual switching rates in our study ranged from 0.06% to 0.73%. Limited access to VL monitoring likely explains these low switching rates. When PHIA data were collected, countries mainly relied on CD4 monitoring and clinical monitoring with limited access to targeted VL testing [10,31–33]. A multi-cohort analysis of 32 HIV treatment programs in sub-Saharan Africa showed that the monitoring strategy used by an ART program is the strongest predictor of switching rate [7]. Without routine VL monitoring, many patients with treatment failure were not switched to second-line ART, or patients were switched late and at low CD4 cell counts, which can contribute to development of resistance and the transmission of resistance and can reduce the likelihood of a sustained benefit of second-line ART [7]. Our study demonstrates the limitations of clinical and immunological monitoring of ART and supports country-level efforts to scale-up VL testing for early detection of NVL. In recent years, southern African countries have made substantial progress in scaling-up laboratory capacity for VL testing, which should improve the capacity of ART programs to identify patients with NVL [7]. In settings in which VL testing is limited, CD4-based targeted VL monitoring may be a useful strategy for monitoring ART [34–36].

Once patients with NVL have been identified, health care providers need to distinguish between patients who require adherence support and those who need a new ART regimen. We noted that more than one-quarter of patients tested negative for recent exposure to ARVs, indicating that a temporary interruption of ART was the likely reason for NVL in these patients. Young women, those with high CD4 cell counts, and those who were on ART for <2 years were more likely to interrupt ART. Suboptimal retention and adherence among women who started ART during pregnancy regardless of immunological or clinical criteria under Option B+ guidelines may explain these findings [37–39]. In line with previous data, our study suggests that a substantial proportion of patients with NVL interrupted ART and may benefit from adherence interventions. The ANRS 12110 trial showed that one-third of patients with NVL had no viral resistance [40]. The DART trial showed that one-third of patients with NVL re-suppressed without changing their ART regimen [41]. A systematic review of five observational studies from South Africa, Eswatini, Thailand, and France showed that more than 70% of patients with NVL re-suppressed following adherence intervention [42]. In agreement with earlier reports, our study supports WHO guidelines to offer adherence support and confirmatory VL testing to patients with a first NVL to avoid unnecessary switching of ART [9]. Our study also underlines the importance of adherence support for patients who started ART with high CD4 cell counts especially during early treatment.

Optimizing ART regimens likely can support countries’ efforts to increase VLS rates among patients receiving ART. In December 2018, WHO recommended dolutegravir (DTG), tenofovir (TDF), and either lamivudine or emtricitabine (XTC) as the preferred first-line regimen, and low- and middle-income countries are transitioning from the current first-line regimen TDF/XTC/EFV to the new DTG-based regimen [43]. DTG is safer and has a higher genetic barrier than EFV. Data from randomized controlled trials support switching patients with undetectable VL from an EFV-based regimen to a DTG-based regimen; however, currently no evidence supports this switch for patients with detectable or unknown VL [44]. Our finding that about 20% of patients receiving ART had a detectable VL >40 copies/mL underlines the need for data to support switching from a failing current first-line treatment to effective second-line treatments. However, the effectiveness of switching from TDF/XTC/EFV to TDF/XTC/DTG needs to be evaluated. Our data suggest that men, individuals who never married, and those with a poor immunologic response to long-term ART are at increased risk of VF and should be monitored carefully after switching to DTG.

The national representativeness of our findings is an important strength of our study. Estimates for NVL prevalence and rates for switching to second-line ART usually come from randomized controlled trials and research cohorts that are not representative of national ART programs. The richness of PHIA data is a further strength of our study. While most HIV cohort studies rely on limited data collected during routine HIV care, PHIA collects detailed data on a wide range of sociodemographic factors, health-related attitudes, and behaviors, enabling us to explore associations between these factors and HIV-related biomarkers. The large sample size is a further strength of our study. We pooled data from five national surveys to obtain a sample size that was large enough to perform adequately powered multivariable analyses.

Our study has several limitations. Our definition of VF did not rely on drug resistance data, and our ability to distinguish between patients with NVL due to inadequate adherence and patients receiving ineffective ART regimens was limited. We classified patients as having interrupted or failing ART based on self-report of current ART use and laboratory testing for detectable concentrations of selected ARVs. Both data sources have limitations that may lead to misclassification of patients. Self-reported data on ART might be susceptible to recall and social desirability biases, and over-reporting of ART use would lead to misclassification of patients as receiving ART and not having VLS. Likewise, laboratory errors could result in misclassification of patients. The rates of switching to second-line ART in Lesotho, Eswatini, and Zimbabwe may be underestimated because these countries tested only for LPV but not for ATV, although both drugs are WHO-recommended second-line treatments [9]. However, the ARVs tested were selected according to national treatment guidelines, and we assumed very few people in Lesotho, Eswatini, and Zimbabwe would have received ATV. Finally, use to the cross-sectional design of the PHIA survey, we could not examine changes in virologic outcomes over the course of ART and had no data on characteristics of patients at the start of first-line or second-line ART.

## Conclusions

The five countries included in our analysis have low rates of NVL and are approaching UNAIDS VLS targets, despite limited access to VL monitoring and possible suboptimal clinical management of patients with NVL. Treatment support for asymptomatic HIV-positive patients initiating ART, scale-up of VL monitoring, and optimized ART regimens may further reduce the prevalence of NVL and may help countries achieve the UNAIDS target of 95% of patients receiving ART having VLS.

## Data Availability

Public datasets for Eswatini, Malawi, and Zambia are available. Public datasets for Lesotho and Zimbabwe will be made available soon. For more information see:  
https://phia-data.icap.columbia.edu/

## Authors’ contributions

AH, ER, and JJ conceptualized the study. AH did statistical analysis and wrote the first draft of the manuscript which was revised by ER and JJ. All authors contributed to interpretation of data and provided critical inputs in the draft manuscript. ER, HA, AJ, NP, SJ, SS, AL, HP, AS, JR, KF, EK, GB, DW, BP, KS, DB, TK, SB, GM, OM, BB, KS, LM, KT, TA, KB, AV, and JJ contributed to design and implementation of PHIA surveys. All authors have read and approved the final manuscript.

## Acknowledgments

PHIA is conducted under the leadership of the respective countries’ Ministries of Health, U.S. Centers for Disease Control and Prevention (CDC), and ICAP at Columbia University (http://www.icap.columbia.edu/).

Collaborating institutions: Eswatini: Ministry of Health; CDC; Central Statistical Office. Lesotho: Ministry of Health; CDC; Bureau of Statistics; National University of Lesotho. Malawi: Ministry of Health; CDC; Centre for Social Research; Chancellor College, University of Malawi; National AIDS Commission; National Statistics Office; Blantyre Health Research and Training Trust. Zambia: Ministry of Health; CDC; Central Statistical Office; Tropical Diseases Research Centre University of Zambia; University Teaching Hospital; National HIV/AIDS/STI/TB Council; Zambian National Public Health Institute. Zimbabwe: Ministry of Health and Child Care, CDC; National Statistics Agency; the Biomedical Research and Training Institute; Lancet Laboratories. South Africa: University of Cape Town (UCT). USA: ICAP at Columbia University; CDC Atlanta; Johns Hopkins University Laboratories Statistical Center for HIV/AIDS Research and Prevention; WESTAT.

## Disclaimer

The findings and conclusions in this report are those of the authors and do not necessarily represent the official position of the funding agencies.

## Supplementary appendix

### Text S1: Laboratory methods for processing of HIV-positive specimens

All HIV-positive results were laboratory-confirmed using Bio-Rad Geenius HIV1/2 (Bio-Rad, Hercules, CA). People who tested HIV-seropositive received PIMA point-of-care CD4 testing (Alere, Waltham, MA) with immediate return of results and submitted plasma specimens for HIV-1 RNA VL testing using the Abbott m2000 System (Abbott Molecular Inc., Chicago, IL) or the Roche COBAS AmpliPrep/COBAS TaqMan platform (Roche Molecular Diagnostics, Pleasanton, CA). The same platforms were used to test dried blood spots (DBS) for VL, in addition to the NucliSENSTM EasyQ HIV-1 v2.0 assay on the bioMérieux (Marcy-l’Étoile, France) for Zimbabwe.

A qualitative high-performance liquid chromatography/tandem mass spectrometry assay was performed to detect antiretroviral drug (ARV) in DBS at the Division of Clinical Pharmacology, University of Cape Town. The samples were processed with a protein precipitation extraction method. Deuterated internal standards were used for each analyte. The extraction procedure was followed by liquid chromatographic separation using a Phenomenex Kinetex EVO C18 (Phenomenex Helvetia, Basel, Switzerland) (1.7 μm; 2.1 × 50 mm; 100 Å) analytical column. An AB Sciex API 4000 mass spectrometer (SCIEX, Framingham, MA, USA) at unit resolution in the multiple reaction monitoring mode was used to monitor the transition of the protonated precursor ions m/z 705.6, 316.0, 629.6, and 267.1 to the product ions m/z 168.2, 243.9, 447.3, and 226.0 for atazanavir, efavirenz, lopinavir, and nevirapine respectively. Electro Spray ionisation was used for ion production. The assay was validated over the range 0.02–5.0 μg/mL, and 0.02 μg/mL was used as the cut-off concentration.

### Text S2: Ethical oversight over the Population-based HIV Impact Assessment (PHIA) surveys in participating countries

The Eswatini Scientific and Ethics Committee, the National Health Science Research Committee Malawi, the National Health Research Ethics Committee Lesotho, the National Health Research Ethics Committee Lesotho, the Tropical Diseases Research Centre Ethics Review Committee, Zambia, the Medical Research Council of Zimbabwe, and the Institutional Review Boards at the Centers for Disease Control and Prevention (CDC; Atlanta, GA) and Columbia University Medical Center (New York, NY) approved the PHIA surveys.

**Table S1:** Characteristics of HIV status disclosure among adults on antiretroviral therapy who participated in the Population-based HIV Impact Assessment (PHIA) survey in Lesotho, Malawi, Eswatini, Zambia and Zimbabwe (2015–2017).

|  | Lesotho | Malawi | Eswatini | Zambia | Zimbabwe | Total |
| --- | --- | --- | --- | --- | --- | --- |
|  | N=2,177 | N=1,406 | N=2,025 | N=1,424 | N=2,168 | N=9,200 |
| Self-reported HIV status, n (%) | 2,177<br>(100.0) | 1,406<br>(100.0) | 2,025<br>(100.0) | 1,424<br>(100.0) | 2,168<br>(100.0) | 9,200<br>(100.0) |
| Positive | 2,065<br>(94.1) | 1,321<br>(93.6) | 1,967<br>(96.7) | 1,296<br>(90.7) | 2,051<br>(93.7) | 8,700<br>(93.1) |
| Negative | 61 (3.3) | 51 (3.6) | 45 (2.6) | 81 (5.7) | 81 (4.5) | 319 (4.3) |
| Never tested/unknown/refused | 51 (2.7) | 34 (2.9) | 13 (0.7) | 47 (3.6) | 36 (1.8) | 181 (2.5) |
| Disclosure to no one, n (%) | 2,065<br>(100.0) | 1,321<br>(100.0) | 1,967<br>(100.0) | 1,296<br>(100.0) | 2,051<br>(100.0) | 8,700<br>(100.0) |
| No | 1,997<br>(96.7) | 1,292<br>(97.6) | 1,910<br>(97.1) | 1,262<br>(97.7) | 2,002<br>(97.7) | 8,463<br>(97.5) |
| Yes | 68 (3.3) | 29 (2.4) | 57 (2.9) | 34 (2.3) | 49 (2.3) | 237 (2.5) |
| Disclosure to spouse/sex partner, n (%) | 2,065<br>(100.0) | 1,321<br>(100.0) | 1,967<br>(100.0) | 1,296<br>(100.0) | 2,051<br>(100.0) | 8,700<br>(100.0) |
| No | 1,007<br>(47.7) | 521 (40.2) | 1,045<br>(52.9) | 525 (39.8) | 824 (38.9) | 3,922<br>(41.2) |
| Yes | 1,058<br>(52.3) | 800 (59.8) | 922 (47.1) | 771 (60.2) | 1,227<br>(61.1) | 4,778<br>(58.8) |
| Disclosure to a doctor, n (%) | 2,065<br>(100.0) | 1,321<br>(100.0) | 1,967<br>(100.0) | 1,296<br>(100.0) | 2,051<br>(100.0) | 8,700<br>(100.0) |
| No | 1,759<br>(84.3) | 1,001<br>(75.7) | 1,898<br>(96.5) | 1,050<br>(80.0) | 1,664<br>(82.3) | 7,372<br>(81.1) |
| Yes | 306 (15.7) | 320 (24.3) | 69 (3.5) | 246 (20.0) | 387 (17.7) | 1,328<br>(18.9) |
| Disclosure to a friend, n (%) | 2,065<br>(100.0) | 1,321<br>(100.0) | 1,967<br>(100.0) | 1,296<br>(100.0) | 2,051<br>(100.0) | 8,700<br>(100.0) |
| No | 1,617<br>(77.7) | 1,095<br>(83.5) | 1,885<br>(95.6) | 1,098<br>(84.2) | 1,696<br>(84.0) | 7,391<br>(84.1) |
| Yes | 448 (22.3) | 226 (16.5) | 82 (4.4) | 198 (15.8) | 355 (16.0) | 1,309<br>(15.9) |
| Disclosure to a family member, n (%) | 2,065<br>(100.0) | 1,321<br>(100.0) | 1,967<br>(100.0) | 1,296<br>(100.0) | 2,051<br>(100.0) | 8,700<br>(100.0) |
| No | 540 (27.5) | 344 (25.8) | 660 (33.7) | 291 (22.6) | 507 (26.5) | 2,342<br>(26.0) |
| Yes | 1,525<br>(72.5) | 977 (74.2) | 1,307<br>(66.3) | 1,005<br>(77.4) | 1,544<br>(73.5) | 6,358<br>(74.0) |

Abbreviations: N, Number of eligible participants; n, number of participants providing a valid response (weighted %).

**Table S2:** Prevalence of alcohol use disorder among adults on antiretroviral therapy who participated in the Population-based HIV Impact Assessment (PHIA) survey in Malawi, Eswatini, Zambia, and Zimbabwe (2015–2017).

|  | Total |
| --- | --- |
|  | N=7,023 |
| AUDIT-C screening outcome, n (%) | 6,915 (100.0) |
| Negative | 6,418 (91.7) |
| Positive | 497 (8.3) |
| AUDIT-C score, n (%) | 6,915 (100.0) |
| 0 | 5,691 (80.1) |
| 1-2 | 573 (9.1) |
| 3-4 | 359 (5.6) |
| 5-8 | 248 (4.4) |
| 9-12 | 44 (0.8) |

Based on Alcohol Use Disorder Identification scale (AUDIT-C)
Abbreviations: N, Number of eligible participants; n, number of participants providing a valid response (weighted %).
In men, an AUDIT-C score of $\geq 4$ and in women, a score of $\geq 3$ is considered positive for hazardous drinking or active alcohol use disorder. Lesotho was excluded from this analysis because AUDIT-C was not collected in this country.

**Table S3:** Sensitivity analysis of rates of switching to second-line ART among HIV-positive adults who participated in the Population-based HIV Impact Assessment (PHIA) survey in Lesotho, Malawi, Eswatini, Zambia, and Zimbabwe (2015–2017).

| Country | Assumed timing of switching |  |  |
| --- | --- | --- | --- |
|  | 1.85 years after ART initiation | 6 months after ART initiation | Day of survey |
|  | Annual rate in %<br>(95% CI) | Annual rate in %<br>(95% CI) | Annual rate in %<br>(95% CI) |
| Lesotho | 0.22 (0.14-0.36) | 0.22 (0.14-0.36) | 0.22 (0.14-0.36) |
| Malawi | 0.15 (0.08-0.35) | 0.15 (0.08-0.35) | 0.15 (0.08-0.35) |
| Eswatini | 0.72 (0.54-0.96) | 0.72 (0.55-0.97) | 0.69 (0.53-0.92) |
| Zambia | 0.73 (0.52-1.04) | 0.74 (0.53-1.05) | 0.70 (0.51-0.99) |
| Zimbabwe | 0.06 (0.02-0.33) | 0.06 (0.02-0.34) | 0.06 (0.02-0.33) |
| **Total** | 0.31 (0.25-0.40) | 0.31 (0.25-0.40) | 0.31 (0.24-0.39) |

Data are annual rates of switching to second-line antiretroviral therapy (ART) in percent and 95% confidence intervals (CI).

